# Global, regional, and national burden and inequalities of infective endocarditis, 1990–2021: A comprehensive analysis from the Global Burden of Disease Study 2021

**DOI:** 10.1101/2025.05.13.25327568

**Authors:** Zhichen Dong, Xiaoli Liu, Yan Zhang

## Abstract

**Aims:** To comprehensively assess the global, regional, and national burden of infective endocarditis (IE) from 1990 to 2021, explore socioeconomic inequalities, identify performance gaps across countries, and project future trends through 2036.

**Methods:** We extracted IE-related data from the Global Burden of Disease Study 2021, including incidence, prevalence, mortality, and disability-adjusted life years (DALYs). Age-standardized rates (ASR) were analyzed across sex, time, and sociodemographic index (SDI) levels. Inequality was quantified using slope and concentration indices. Frontier analysis assessed performance gaps across countries. Decomposition analysis explored contributors to DALY changes, and future trends were projected using a Bayesian age-period-cohort model.

**Results:** Between 1990 and 2021, global DALYs due to IE increased by 55.67%, mainly driven by population growth (87.92%) and aging (38.91%), while the DALY ASR declined by 9.75%. Marked regional and socioeconomic disparities were observed: high-SDI regions showed rising incidence and prevalence, while low- and middle-SDI regions exhibited decreasing ASRs but persistent burdens. Socioeconomic inequality in IE burden has narrowed over time, yet remains significant. Epidemiological transitions contributed to reduced burden in some settings, but structural disparities continue to affect outcomes. Projections indicate further increases in incidence and prevalence through 2036, despite ongoing declines in deaths and DALYs.

**Conclusion:** IE remains a growing global health challenge with persistent inequalities. While overall burden has shifted, socioeconomic disparities continue to shape disease patterns. These findings highlight the need for equity-focused, context-specific strategies to reduce the global burden of IE.

**Lay Summary:** This study explores how the global burden of infective endocarditis has changed over time and highlights persistent inequalities across countries with different development levels.

**Key findings:**

• Although the age-standardized rate of IE has declined globally, the total number of cases and deaths has risen, mainly due to population growth and aging.
• Socioeconomic inequality in IE burden has decreased over time but remains substantial, underscoring the need for more equitable healthcare interventions worldwide.

## Introduction

Infective endocarditis (IE) is a life-threatening infectious disease characterized by microbial colonization of cardiac valves or endocardial tissue, posing a significant global health and economic burden. ^1–4^ In 2019, IE accounted for over 1,090.5 thousand incident cases and 1,723.5 thousand disability-adjusted life years (DALYs) worldwide, reflecting substantial health and economic tolls. ^5–7^ Despite advancements in diagnostics and therapeutics, mortality remains alarmingly high, with in-hospital mortality over 25%, and 5-year mortality over 40%^8–12^. Septic shock, vascular embolism, and valve destruction are important causes of IE death. ^13^

Globally, IE is experiencing an epidemiological transition driven by demographic changes and evolving clinical practices. ^4^ Aging populations, the rising prevalence of chronic comorbidities (e.g., diabetes and end-stage renal disease requiring hemodialysis), and increased utilization of invasive cardiac interventions have altered the incidence patterns and risk profiles of IE worldwide. ^14,15^ Notably, the rapid expansion of valve replacement procedures and escalating antimicrobial resistance have further complicated IE management. ^3,16,17^ Moreover, recent trends highlight intravenous drug use (IVDU) as an emerging contributor, particularly among younger populations. For instance, hospitalizations related to IVDU-associated IE among individuals aged 15–34 years nearly doubled from 6.9% in 2005 to 12.1% in 2016. ^18,19^

Significant disparities exist across socioeconomic regions. Low- and middle-income countries disproportionately experience complications such as heart failure and persistent infection due to limited healthcare access. ^20,21^ Rheumatic heart disease (RHD) remains the predominant underlying etiology in low-income areas, associated with approximately half of IE cases. ^21,22^ In contrast, degenerative valvular diseases and congenital heart diseases have supplanted RHD as primary risk factor in high-income countries and territories, driven largely by aging populations and improved cardiovascular care. ^23^

Although several studies have investigated the global burden of IE, they relied on older data, shorter observation periods, or region-specific analyses. ^5–7,24,25^ Critically, prior research lacks in-depth exploration of socioeconomic and healthcare inequalities in IE epidemiology across regions and nations. Due to the high mortality rate and ongoing epidemiological shifts of IE, understanding its epidemiology, regional differences, and health inequalities is crucial for guiding equitable resource allocation and health policy development.

Utilizing data from the Global Burden of Disease Study (GBD) 2021, this study presents an updated and comprehensive evaluation of the burden, temporal trends, and healthcare inequalities associated with IE. Specifically, it describes the temporal trends of IE at global, regional, and national levels, assesses inequalities across countries, investigates demographic and epidemiological factors through decomposition analyses, and projects the global burden of IE up to the year 2036.

## Methods

### Data Sources

The GBD 2021 study conducted a comprehensive assessment of health losses related to 369 diseases, injuries, and impairments, as well as 88 risk factors across 204 countries and territories, utilizing updated epidemiological data and refined standardization methodologies.^26,27^ Data collated in the GBD database were modeled through spatiotemporal Gaussian process regression to enable age-, time-, and location-specific smoothing in regions with incomplete datasets. A Bayesian prior-regularized and trimmed Meta-Regression (MR-BRT) framework was employed to adjust for data biases arising from heterogeneous case definitions and research methodologies across nations. These analytical approaches have been extensively detailed elsewhere. For the GBD 2021 evaluation, IE was clinically defined as per prior studies, with ICD-10 codes (I33-I33.9, I38-I39.9) and ICD-9 codes (074.22, 421-421.9) used for case identification in vital registration systems. The Institutional Review Board of Peking University First Hospital exempted this study from ethical approval due to its reliance on publicly available data. This research adheres to the Guidelines for Accurate and Transparent Health Estimates Reporting (GATHER).^28^ Sociodemographic index The Sociodemographic Index (SDI) is a composite metric integrating total fertility rate, per capita income, and educational attainment, serving as a proxy for societal development ranging from 0 to 1.^26,27^ Higher SDI values indicate more advanced socioeconomic conditions. In this study, 204 countries and territories were stratified into five SDI-based quintiles (low, low-middle, middle, high-middle, and high) to investigate associations between IE burden and socioeconomic development.

### Global Burden and Trend Assessment

This study evaluated the global burden of IE through case numbers, age-standardized rates (ASR) and average annual percentage changes (AAPC) of incidence, prevalence, deaths and DALYs. ASR per 100,000 population enabled cross-regional comparisons of disease burden across heterogeneous age structures, while AAPC with 95% confidence intervals (CI) served as composite indicators reflecting temporal trends in ASR. AAPC, which is a summary measure of trends over a defined period, was calculated via 32-year trend analysis using Joinpoint software (version 5.3.0.0). This tool segments overall trends into multiple sub-periods by detecting inflection points (up to 6 joinpoints forming 7 final segments) to identify localized temporal patterns. Smoothing spline models visualized dynamic relationships between age-standardized disease burden metrics and the Sociodemographic Index (SDI) across 204 countries in 2021, as well as 21 GBD regional trends during 1990-2021, with Spearman’s rank correlation quantifying associations.

### Cross-country Inequality Analysis

To assess regional inequalities in disease burden, we applied the slope index of inequality (SII) and concentration index (CI) to evaluate DALY disparities. SII is calculated by regressing DALYs rates across countries against their relative positions on the sociodemographic development spectrum, reflecting absolute disparities in disease burden between countries with the highest and lowest levels of economic development. CI quantifies relative cross-national health inequalities by measuring deviations between the cumulative fraction of DALYs and cumulative population ranked by SDI through the Lorenz concentration curve. For both SII and CI, positive values indicate a heavier burden in high-SDI countries, while negative values signify greater burden in low-SDI countries.

### Frontier analysis

We utilized frontier analysis to identify effective disparities between observed DALY rates and the minimum achievable burden for each country’s development level. This approach quantified maximum potential DALY reductions across the SDI spectrum by determining feasible age-standardized DALY rates at given developmental stages. The magnitude of effective disparity from the frontier quantifies a nation’s untapped potential for DALY improvement relative to its SDI position.

### Decomposition Analysis

To elucidate the explanatory factors driving changes in IE DALYs from 1990 to 2021, we conducted a decomposition analysis stratified by population size, age structure, and epidemiological shifts. The DALY count for each location was calculated as the formula: 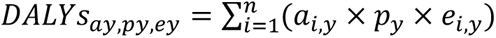, where DALYs_ay, py, ey_ represents the total DALYs according to the factors of age structure, population size, and DALYs rate in year y; a_i,y_ denotes the population proportion of the i-th age group among n age strata in year y; p_y_ denotes the total population in year y; and e_i,y_ the DALY rate of the i-th age group in year y^29^. The contribution of each factor to DALY changes over 1990–2021 was quantified by isolating its temporal variation while holding other factors constant.

### Predictive Analysis

To guide evidence-based public health strategizing and optimize healthcare resource allocation in the future, we extended our analysis to project the future burden of IE through 2036. This prospective modeling leveraged an integrated Bayesian Age-Period-Cohort (BAPC) framework coupled with the Integrated Nested Laplace Approximation (INLA) algorithm, a computational advance demonstrating enhanced predictive reliability through superior credible interval coverage and reduced estimation error compared to conventional age-period-cohort (APC) methodologies. The BAPC-INLA synthesis rigorously accounts for temporal fluctuations in disease surveillance, generational risk exposure variations, and period-specific therapeutic advancements, thereby generating robust global projections that inform priority-setting in cardiovascular infection control.

## Results

### Global trends

Globally, the number of incident IE cases increased by 135.0% from 1990 to 2021, reaching 1,042,477.45 (95% UI: 893,665.11–1,204,150.02), with an AAPC of 0.97 (95% CI: 0.94–1.00) (Figure 1 A-B, Supplementary Table S1). Prevalent cases grew more rapidly, rising 200.0% from 140,559.09 (95% UI: 121,657.59–161,472.54) in 1990 to 421,667.22 (95% UI: 362,727.60–482,472.13) in 2021. The global ASR of prevalence increased at an AAPC of 1.93 (95% CI: 1.83– 2.03) over the 32-year period (Figure 1 C-D, Supplementary Table S2). For mortality, the crude death count rose from 36,883.49 (95% UI: 31,645.96–40,522.30) in 1990 to 77,843.89 (95% UI: 69,010.02–86,337.76) in 2021, while the ASR showed a modest decline (AAPC -0.02, 95% CI: -0.13–0.10) (Figure 1 E-F, Supplementary Table S3). Global DALYs in 2021 were estimated at 2,076,412.89 (95% UI: 1,827,083.81–2,308,504.19), with an ASR of 25.56 (95% UI: 22.34–28.37) and an AAPC of -0.33 (95% CI: -0.43 to -0.23) (Figure 1 G-H, Supplementary Table S4). Global trends by sex From 1990 to 2021, males consistently exhibited higher ASRs than females across incidence, prevalence, deaths, and DALYs. Both incidence and prevalence rates showed continuous increases over the 32-year period in all groups, with males experiencing the steepest rises. In contrast, ASRs of deaths and DALYs peaked around 2001, followed by a gradual decline, more pronounced in females (Figure 2).

**Figure 1.**
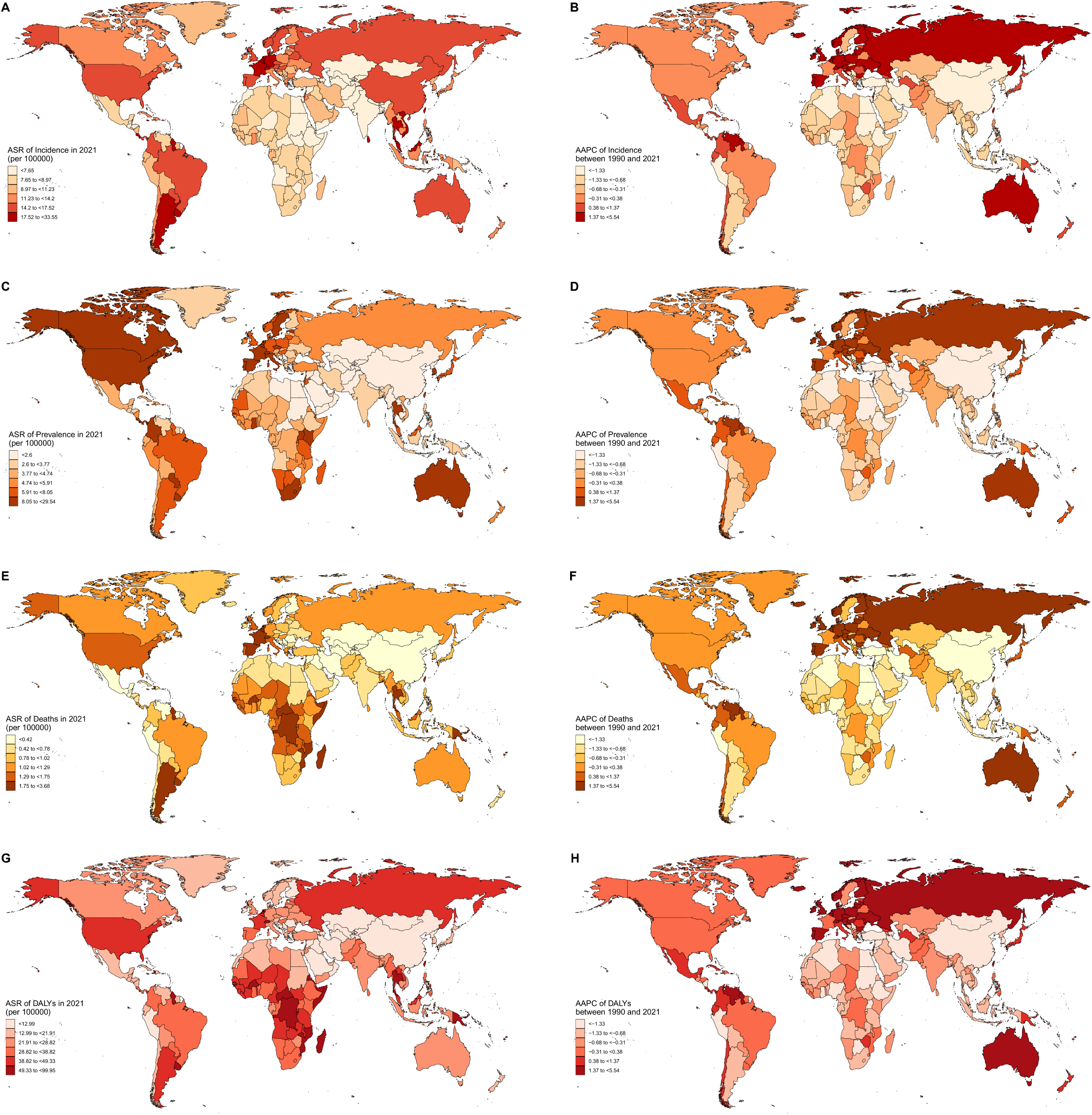
The global disease burden and temporal trends of infective endocarditis for both sexes in 204 countries and territories. (A) The ASR of incidence in 2021; (B) The trend in ASR of incidence (AAPC) from 1990 to 2021; (C) The ASR of prevalence in 2021; (D) The trend in ASR of prevalence (AAPC) from 1990 to 2021; (E) The ASR of deaths in 2021; (F) The trend in ASR of deaths (AAPC) from 1990 to 2021; (G) The ASR of DALYs in 2021; (H) The trend in ASR of DALYs (AAPC) from 1990 to 2021. AAPC, average annual percentage change; ASR, age-standardized rate; DALYs, disability-adjusted life-years.

**Figure 2.**
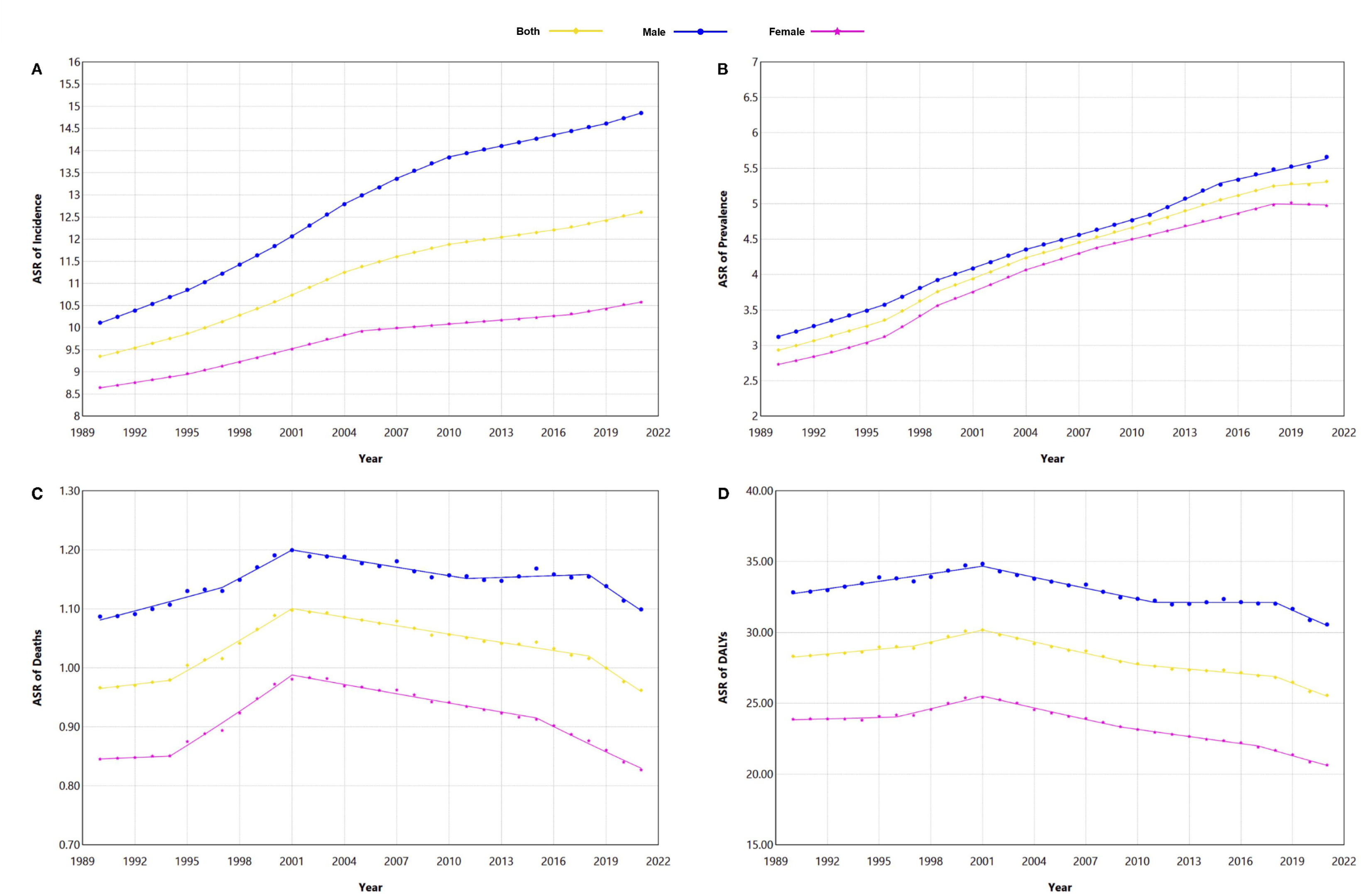
Joinpoint regression analysis of age-standardized rates of global infective endocarditis for both sexes from 1990 to 2021. (A) Joinpoint regression analysis of age-standardized incidence rate; (B) Joinpoint regression analysis of age-standardized prevalence rate; (C) Joinpoint regression analysis of age-standardized death rate; (D) Joinpoint regression analysis of age-standardized Incidence rate.

### Regional and national trends

The global burden of IE demonstrates significant regional disparities associated with SDI. A positive correlation exists between SDI and ASR of incidence (R=0.62, p<0.001) or prevalence (R=0.45, p<0.001) (Supplementary Figure S1). The high-SDI quintile had the highest ASR of incidence (15.77, 95% UI: 13.63–18.08) and ASR of prevalence (9.81, 95% UI: 8.49–11.17), while the low-SDI quintile showed the lowest ASR of incidence (7.18, 95% UI: 6.34–8.30), and the low-middle SDI quintile the lowest ASR of prevalence (3.32, 95% UI: 2.81–3.92) (Supplementary Table S1-2). Mortality and DALYs exhibited U-shaped SDI relationships: the middle-SDI quintile had the lowest ASR of deaths (0.56, 95% UI: 0.49–0.71) and ASR of DALYs (17.93, 95% UI: 15.87–22.40), whereas the high-SDI quintile showed the highest ASR of deaths (1.34, 95% UI: 1.18–1.44), and low-SDI quantile showed the highest ASR of DALYs (40.71, 27.37–52.99) (Supplementary Table S3-4). Notably, the high-SDI quintile was the only group with rising ASR of DALYs.

Geographically, Southern Latin America had the highest ASR of incidence (18.36, 95% UI: 16.14–20.59) and fastest growth (AAPC 1.45, 95% CI: 1.40–1.51). Oceania led in ASR of deaths (2.14, 95% UI: 1.57–3.04) and ASR of DALYs (75.41, 95% UI: 54.52–104.32). Western Europe, High-income North America, and Australasia ranked highest in ASR of prevalence. Eastern Europe showed the most rapid increases in ASR of incidence (AAPC 2.05, 95% CI: 1.99–2.10) and ASR of DALYs (AAPC 4.06, 95% CI: 2.28–5.86). Central Asia (ASR of DALYs: 6.39, 95% UI: 5.57–7.29) and East Asia (ASR of DALYs: 4.59, 95% UI: 3.72–6.22) exhibited the lightest burdens, with equally pronounced declines. In Central and Eastern Sub-Saharan Africa, despite low ASR of incidence, ASR of deaths and DALYs remained persistently elevated (Supplementary Table S1-4, Figure S3-6).

Global IE burden across 204 countries and territories revealed stark disparities. Thailand had the highest ASR of incidence (33.55, 95% UI: 29.76–37.89), while Western Europe dominated prevalence and mortality: France (ASR of prevalence: 29.54, 24.76–35.05) and Switzerland (ASR of deaths: 3.68, 2.98–4.11) led their respective metrics, with the Netherlands ranking second in both. Small island nations bore the heaviest DALYs: Tokelau (99.95, 76.93–135.08), Madagascar (91.07, 56.51–135.06), and the Marshall Islands (88.55, 58.87–126.01). Conversely, Central Asian countries showed minimal burden: Tajikistan had the lowest ASR of incidence (4.31, 3.52–5.27), while Azerbaijan reported the lowest ASR of prevalence (0.46, 0.39–0.55), deaths (0.03, 0.02– 0.04), and DALYs (1.09, 0.70–1.63), reflecting regional success in rheumatic fever control (Supplementary Table S5).

Temporal trends (1990–2021) highlighted Singapore’s fastest incidence growth (AAPC 2.82, 95% CI: 2.68–2.96) and Taiwan (China)’s steepest prevalence rise (AAPC 6.56, 6.15–6.96). Taiwan (China), Russia, and Guyana led death and DALY growth. (Supplementary Table S7-8) Declines clustered geographically: Nigeria (-0.76, -0.82–-0.70) and Liberia in Western Sub-Saharan Africa reduced incidence; Uzbekistan (-0.64, -0.88–-0.40) and Kazakhstan in Central Asia lowered prevalence. China (-4.09, -4.32–-3.86 for deaths; -4.86, -5.16–-4.56 for DALYs) and South Korea in East Asia achieved the sharpest death and DALY reductions (Supplementary Table S6).

### Cross-country inequality analysis

Between 1990 and 2021, both absolute and relative inequalities in DALY burden across countries showed noticeable changes. In 1990, the slope index of inequality (SII) was -21.7 (95% CI: -27.5 to -18.3), indicating a substantially higher burden in lower-SDI countries. By 2021, the SII increased to 5.04 (95% CI: -3.59 to 13.66), reflecting a reversal in the direction of inequality, though the estimate was not statistically significant (Figure 3 A). The concentration index (CI) also shifted over time. In 1990, CI was -0.029 (95% CI: -0.508 to 0.558), suggesting a slight and statistically non-significant concentration of burden among countries with lower SDI. In 2021, the CI rose to 0.079 (95% CI: -0.036 to 0.624), indicating a mild redistribution of DALYs toward countries with higher SDI levels (Figure 3 B).

**Figure 3.**
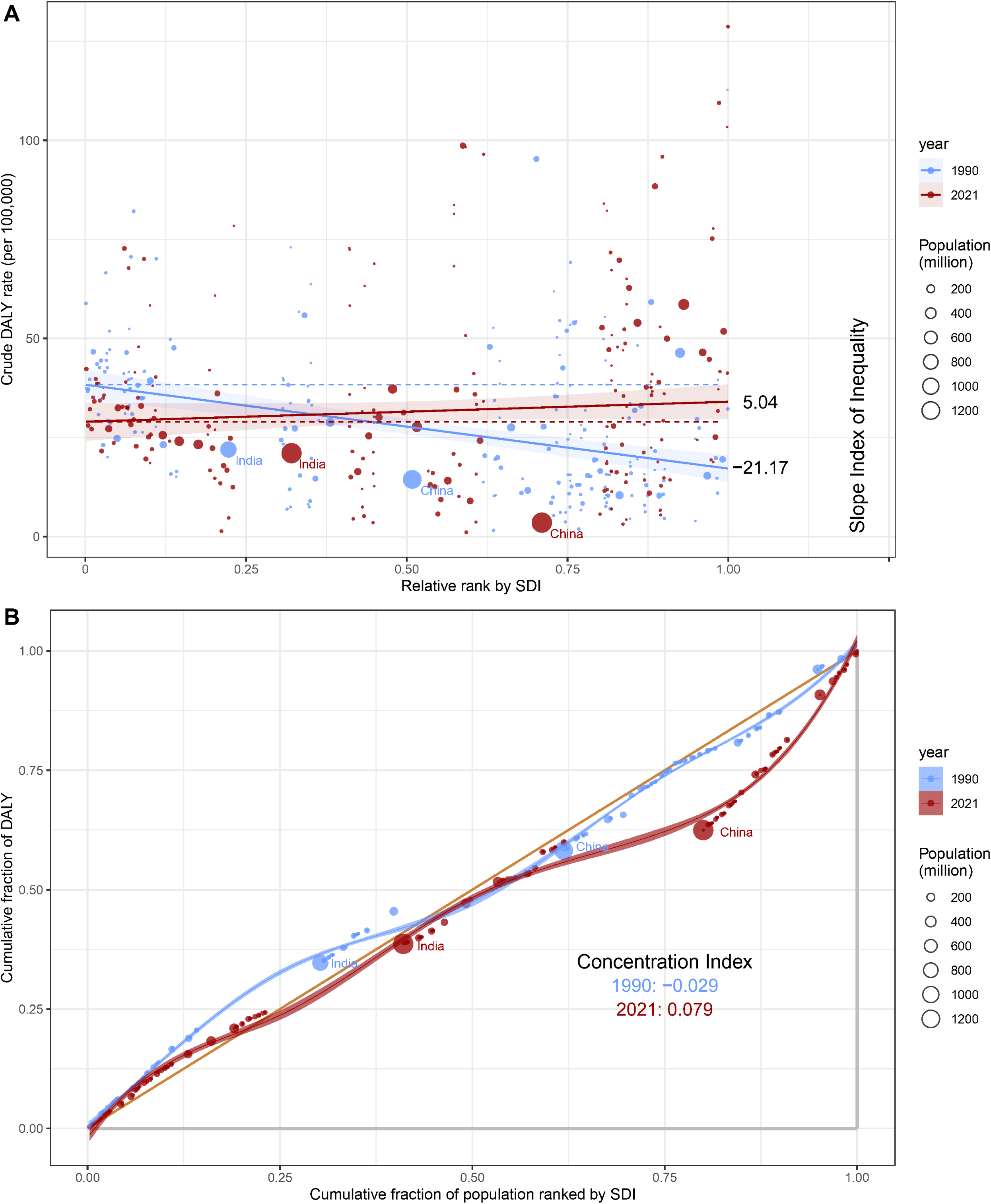
Socio-demographic inequality in the DALY burden of infective endocarditis: (A) regression-based slope index of inequality and (B) concentration curves, 1990 vs. 2021. DALY, disability-adjusted life year.

### Frontier analysis of age-standardized DALY rates

While higher-SDI nations generally exhibited smaller effective disparities, our analysis revealed that developed countries were not the sole strong performers. For instance, low-SDI countries such as Somalia, Niger, and Mali demonstrated minimal effective disparities. Conversely, high-SDI nations like Netherlands, Luxembourg, and Switzerland showed larger effective disparities. Additionally, countries including Tokelau, Madagascar, the Marshall Islands, and Thailand had the heaviest IE-related DALY burdens relative to their SDI levels (Figure 4).

**Figure 4.**
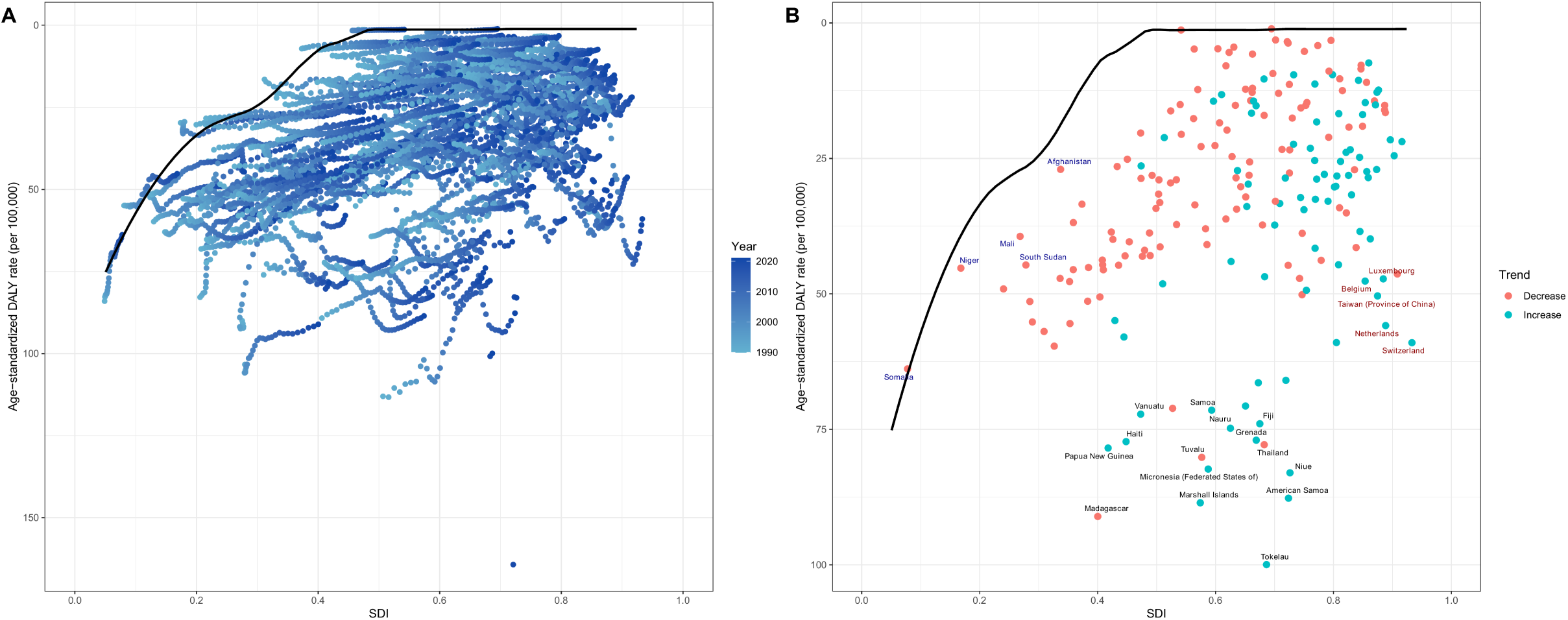
Frontier analysis of DALY burden from infective endocarditis across 204 countries and territories, 2021. (A) Construction of the efficiency frontier curve based on the lowest observed DALY rates by SDI; (B) Distance of each country from the frontier line, representing relative performance gaps in health system efficiency. DALYs, disability-adjusted life years; SDI, sociodemographic index.

### Decomposition analysis on DALYs

From 1990 to 2021, the global burden of IE, measured in DALYs, increased by 55.67%, while the ASR declined by 9.75%, showing heterogeneous growth patterns across SDI regions. All regions except high-SDI areas exhibited declines in ASR to varying degrees. The primary contributor to the global increase in IE burden was population growth (87.92%), followed by aging (38.91%), and epidemiological changes (-26.82%). These patterns were consistent across sexes. Population growth had the most significant impact on lower-SDI regions, with the largest increase in burden observed in low-SDI countries (139.5%). In high-SDI regions, aging emerged as the dominant driver of DALY growth (47.56%). Epidemiological changes contributed negatively to DALY trends globally and across the four SDI quintiles, except for high-SDI regions (23.79%), with the most significant decline observed in middle-SDI regions (-214.81%) (Figure 5, Supplementary Table S9). Decomposition analysis revealed substantial variations in the contributions of these three determinants across different SDI categories and sexes.

**Figure 5.**
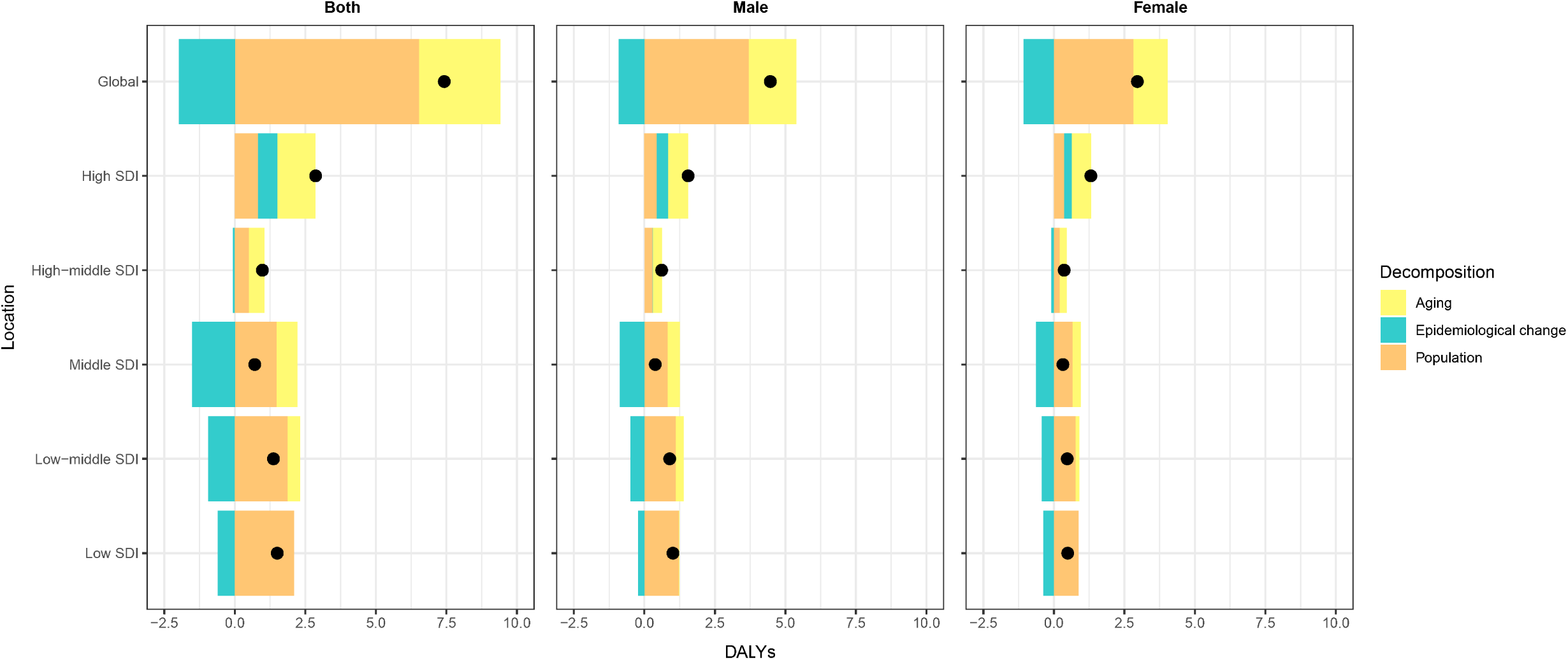
Decomposition of changes in DALYs due to infective endocarditis by sex and SDI quintile, 1990–2021. DALYs, disability-adjusted life-years.

### Predictive analysis on IE burden to 2036

The ASR of incidence will continue to rise for both sexes over the next 15 years, with the overall ASR expected to reach 13.94 per 100,000 population by 2036. In terms of prevalence, sex-specific trends diverge: the ASR of male prevalence is projected to increase steadily, reaching 6.58 per 100,000, whereas the ASR of female prevalence, having peaked in 2022, is expected to decline to 4.85 per 100,000 by 2036. Nonetheless, the overall prevalence ASR is anticipated to follow an upward trajectory, reaching 5.58 per 100,000 by the end of the projection period. In contrast, both the ASR of deaths and the ASR of DALYs are projected to decline from 2022 through 2036 for both males and females. By 2036, the overall ASR of deaths is estimated to decrease to 0.76 per 100,000, while the ASR of DALYs is projected to decline to 20.05 per 100,000. (Figure 6)

**Figure 6.**
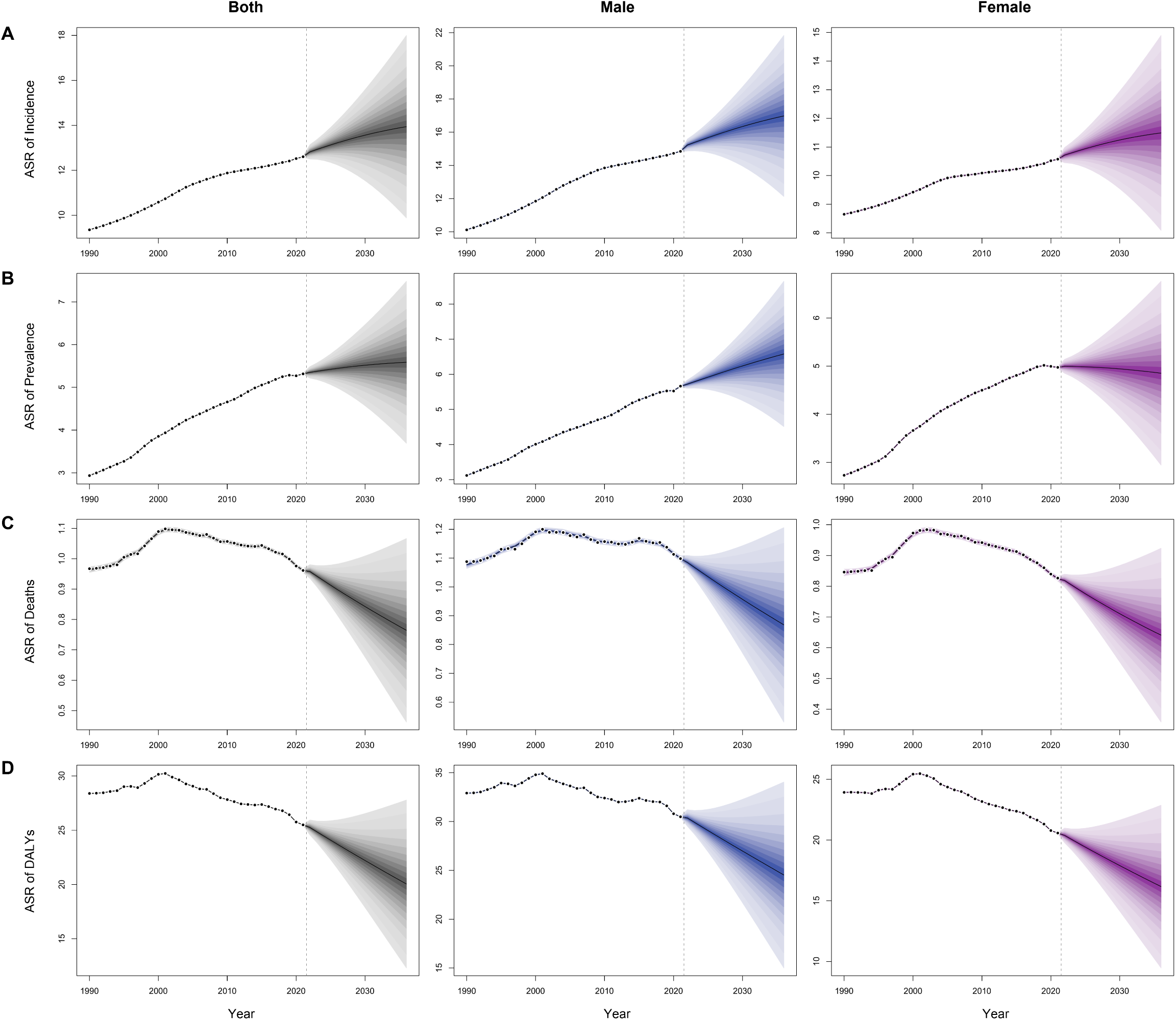
Projected age-standardized rates of incidence (A), prevalence (B), deaths (C), and DALYs (D) for infective endocarditis by sex from 2022 to 2036. DALYs, disability-adjusted life-years.

## Discussion

From 1990 to 2021, the global burden of IE has risen substantially. The number of incident cases and deaths more than doubled over the study period, driven by both population growth and shifts in disease epidemiology. The incidence and prevalence rates have been consistently increasing during the 32 years, while death and DALY rates decreased after 2001. Significant regional differences were observed. High incidence and prevalence are associated with high SDI, while high mortality and DALY are concentrated in areas with the highest and lowest SDI.

Cross-country inequality analysis showed that the inequality associated with SDI is reduced, but the burden of DALYs is shifted from high SDI to low SDI regions. Although death and DALY rates are expected to decrease through 2036, incidence and prevalence will remain on the rise. The control and management of IE will continue to face prominent challenges in the coming decades.

SDI is closely related to IE burden. During the past 32 years, IE burden in high SDI region has significantly aggravated. The ASR of Incidence, prevalence and deaths in high SDI region are the highest, with the rapid growth of IE burden. This is associated with multiple factors. On the one hand, high-SDI regions face severe population aging. Accelerated aging has led to a marked increase in the proportion of individuals aged ≥65 years. ^30^ The prevalence of degenerative valve disease (such as aortic valve calcification) has been increasing with the population ageing, directly leading to an increase in endocardial vulnerability. ^23,31^ Aging will also lead to an increase in chronic diseases such as diabetes and chronic renal disease, becoming a risk of infection and high mortality. ^14,15,32^ On the other hand, widespread use of cardiac implantable devices and invasive medical procedures has heightened iatrogenic infection risks. Permanent pacemakers and defibrillators elevate IE risk by 50%, and IE cases linked to cardiac implantable electronic devices are increasing over time. ^33,34^ Invasive dental treatment and valve replacement procedures have also been important reasons for the increased incidence of IE. ^16,35,36^ In addition, the increasing prevalence of intracardiac device implantation, intravenous drug use and other healthcare exposures has led to a pathogen shift in high-SDI regions. Staphylococcus aureus has replaced Streptococcus viridans as the main pathogen, and multidrug-resistant Staphylococcus aureus (methicillin-resistant, vancomycin-resistant) has resulted in limited options for antibiotic treatment.^37^ In healthcare-advanced regions such as Western Europe, High-income North America and Southern Latin America, these medical procedures are more accessible and prevalent, but also lead to an elevated risk of endocardial infections.

However, due to favorable healthcare-economic conditions, the DALY burden is less prominent in high-SDI regions. In contrast, the regions with the highest DALYs are the less-developed areas including Oceania and Sub-Saharan Africa. In low-SDI regions, the control of rheumatic fever remains inadequate, leading to a high prevalence of RHD. Patients with RHD often suffer from valvular damage, which increases the risk of bacterial adherence. In some areas, poor living conditions, lack of basic sanitation facilities, and limited access to clean water contribute to the spread of infection and bacteremia, increasing the risk of IE. More importantly, the shortage of medical resources — including insufficient diagnostic equipment and a lack of trained healthcare professionals — leads to delays in diagnosis and treatment, as well as inadequate and inappropriate use of antibiotics. As a result, low-SDI regions bear a substantial burden of IE. Given the underdeveloped healthcare systems in some low-SDI areas, data on incidence and mortality may be incomplete, and the actual burden of IE is likely more severe than reported.

A noteworthy finding of this study is that the burden of IE in Central and East Asia has been low and declining over the past three decades. This trend may be attributable to a combination of factors, including the successful control of rheumatic heart disease, expansion of primary healthcare coverage, and effective public health infrastructure. In China, for example, control of rheumatic heart disease, antimicrobial administration, and infrastructure improvements in primary care setting s have likely reduced the incidence of healthcare-associated IE.^38,39^ Additionally, lower rates of intravenous drug use in some countries may contribute to the reduced transmission of IE-related pathogens. The consistent improvement observed in these regions suggests the presence of effective, context-specific strategies that merit further exploration.

Over the past 32 years, the global incidence and prevalence of diseases have been on the rise, which is related to the worsening of global aging, the increase of iatrogenic infections, and the improvement of testing methods in underdeveloped regions. However, there has been a clear turning point in death and DALY trends. Joinpoint regression reveals that the global burden of IE has been on a downward trend since 2001, which is of great significance. The improvement in the burden mainly stems from a milestone in the diagnosis of IE—the Duke criteria, first proposed in 1994 and modified in 2000.^40,41^ The Duke criteria integrate microbiological and imaging evidence and list echocardiographic manifestations as the major criteria for the first time.^40^ Before the Duke criteria, the confirmation of IE relied on pathological evidence during surgery, which led to the failure of many patients to receive an early diagnosis, especially for those with negative blood cultures or who were unable to undergo surgery. The modification of the Duke criteria further emphasize the importance of transesophageal echocardiography and refine the secondary diagnostic criteria, improving sensitivity and specificity.^41^ Through ultrasonic evidence, IE can be quickly identified and intracardiac complications can be detected at an early stage, enabling timely antibiotic or surgical interventions. In addition, the optimization of antibiotic treatment regimens and the progress in cardiac surgery techniques have also contributed to the improvement of the global burden of IE in the 21^st^ century.^42^ The turning point in the trend of DALYs illustrates that timely echocardiography and blood tests play a crucial role in the early diagnosis and treatment of IE. However, echocardiography is still not widely available in many low-SDI areas, which may be an important cause of high mortality in IE.

From 1990 to 2021, the global burden of IE, measured by DALYs, increased by 55.67%, with population growth (87.92%) and aging (38.91%) being the primary drivers. This increase occurred despite a 9.75% decline in the ASR, reflecting overall improvements in mortality. The impact of population growth was most pronounced in low-SDI countries, contributing to a 139.5% rise in DALYs, highlighting the need for enhanced healthcare capacity and access in rapidly growing populations. In high-SDI countries, aging was the leading contributor to the burden (47.56%), consistent with broader epidemiological shifts toward chronic and age-related diseases. Epidemiological changes showed an overall negative contribution to DALY trends globally (-26.82%), with particularly large reductions in middle-SDI countries (-214.81%), suggesting improved prevention, diagnosis, and treatment efforts. However, in high-SDI countries, epidemiological factors contributed positively (23.79%) to DALY growth, likely due to an increasing burden from healthcare-associated infections and intravenous drug use.

Quantifying the transnational inequality of IE burden on SDI gradient can make the burden distribution pattern clearer and identify countries that should improve IE prevention and control. In our research, both the SII and CI indicated significant changes in the distribution of the disease burden. In 1990, the SII revealed a considerably higher burden of IE in countries with lower SDI. However, by 2021, the SII reversed direction, showing a slight increase in DALYs for IE in higher-SDI countries, although the confidence interval for this estimate included zero, suggesting the change was not statistically significant. Similarly, the CI transitioned from a marginally negative value in 1990 to a positive value in 2021, indicating a slight redistribution of the burden toward higher-SDI countries, but the confidence interval again indicated a lack of statistical significance. These changes may reflect significant improvements in the management and prevention of IE in low-SDI and lower-middle SDI countries. With the support of global health initiatives, many low- and middle-income countries have made substantial progress in improving healthcare access, diagnostic technologies, and strengthening health infrastructure.^21^ These advancements may have facilitated early diagnosis and timely treatment, thereby reducing mortality and disease progression. Additionally, public health interventions in these countries, such as improved infectious disease control and better sanitation, may have contributed to alleviating the burden of IE. In contrast, higher-SDI countries may have experienced an increase in the burden due to rising risk factors, such as intravenous drug use and healthcare-associated infections.^23,25^ However, the wide confidence intervals observed for both indices highlight considerable uncertainty regarding the interpretation of these trends, emphasizing the need for further research. Understanding the underlying drivers of these changes in inequality is crucial for developing targeted public health interventions, particularly for reducing the burden of IE in low-SDI and lower-middle SDI countries.

The frontier analysis highlights the disparity between the actual burden of IE and the theoretical optimal values across countries, offering valuable insights for targeted public health interventions. Western European countries like Switzerland, the Netherlands, and Belgium, despite their high SDI, exhibit a relatively high disease burden. These countries share common challenges faced by high-SDI settings, including population aging, antimicrobial resistance, and healthcare-associated infections. For instance, calcific aortic valve disease, often associated with advanced age, poses a significant clinical concern in the Netherlands and Luxembourg.^43^ In Switzerland, MRSA accounts for over 30% of IE cases, highlighting the substantial impact of resistant pathogens on disease burden.^44^ In contrast, countries like Somalia and Niger have achieved relatively low IE burdens despite lower SDI levels, likely due to the lower prevalence of RHD, which is a major risk factor for IE in low-SDI countries.^45^ This suggests that improvements in RHD control and early intervention could significantly reduce the IE burden in these regions. Countries with the heaviest IE burden are predominantly small island nations in Oceania. Given their relatively small populations, these countries exhibit unstable incidence and death rates. Beyond small island nations, Thailand also faces a particularly severe burden of IE. reporting the highest ASR of incidence among all countries globally. The high burden in Thailand may be due to the effects of persistent rheumatic heart disease, aging population, intravenous drug use and zoonotic infections.^46–48^ These findings underscore the need for tailored public health strategies: while high-burden countries require more efficient healthcare systems and better management of risk factors, low-SDI countries benefit from enhanced RHD prevention and early diagnosis programs. In addition, intravenous drug use has affected the incidence of IE in many regions in recent years, such as Eastern Europe and North America.^49–51^ Controlling drug use and related infections is also an important topic for IE prevention.

Projections based on the BAPC-INLA model indicate a continued rise in the ASR of incidence and prevalence over the next 15 years, despite concurrent declines in ASR of deaths and DALYs. The increasing incidence may reflect improved detection, aging populations, and rising exposure to risk factors such as healthcare-associated infections and intravenous drug use. The divergence in prevalence trends between sexes—continued rise in males versus a projected decline in females—suggests potential differences in risk exposure, healthcare access, or disease progression patterns. The projected decline in mortality and DALYs likely reflects advances in diagnosis, treatment, and long-term management of IE. However, the rising overall burden in terms of case numbers and chronic disease prevalence underscores the need for sustained investment in prevention, early detection, and risk factor control.

This study has several limitations. First, the primary data inputs for the GBD model are derived from a wide range of countries, inevitably leading to variations in data quality. This issue is particularly pronounced in underdeveloped regions, where the scarcity of high-quality data may compromise the accuracy of model-based estimates. Second, the underdiagnosis of IE in many developing countries may contribute to an underestimation of the true disease burden. Lastly, temporal comparisons may be affected by changes in diagnostic criteria for IE over the study period, potentially limiting the comparability of results across time.

In conclusion, the global burden of IE increased significantly in absolute terms from 1990 to 2021, mainly driven by population growth and aging, despite declining age-standardized mortality and DALY rates since 2001. Geographic disparities remain evident—high-SDI countries face rising incidence linked to aging and medical interventions, while low-SDI regions struggle with limited resources and prevention efforts. Central and East Asia showed encouraging reductions, highlighting the value of strong health systems. Although inequality has narrowed, frontier analysis reveals ongoing gaps between actual and achievable outcomes. With incidence and prevalence projected to rise further, continued investment in prevention, early detection, and long-term care is essential to reduce the global IE burden.

## Data Availability

The data supporting this study are publicly available in the Global Burden of Disease (GBD) repository at https://vizhub.healthdata.org/

https://vizhub.healthdata.org/gbd-results/

## Acknowledgement

We would like to thank the countless individuals who have contributed to the Global Burden of Disease Study 2021 in various capacities.

## Authors’ Contributions

Zhichen Dong: Data Curation, Formal Analysis, Writing – Original Draft Xiaoli Liu: Conceptualization, Methodology, Writing – Review & Editing Yan Zhang: Conceptualization, Project Administration, Writing – Review & Editing

## Data availability

Data can be accessed through the online query tool available on the Institute for Health Metrics and Evaluation (IHME) website (http://ghdx.healthdata.org/), with no permission required for retrieval.

## Appendices

Supplemental document: Table S1-9 and Figure S1-6.

